# Distribution equality as an optimal epidemic mitigation strategy

**DOI:** 10.1101/2020.09.15.20194506

**Authors:** Adar Hacohen, Reuven Cohen, Sol Efroni, Ido Bachelet, Baruch Barzel

## Abstract

Upon the development of a drug or vaccine, a successful response to a global pandemic, such as COVID-19, requires the capacity for efficient distribution at a global scale. Considering constraints on production and shipping, most existing strategies seek to maximize the outflow of therapeutics, hence optimizing for rapid dissemination. Surprisingly, we find that this intuitive approach is counterproductive. The reason is that focusing strictly on the quantity of disseminated therapeutics, such strategies disregard their specific spreading patterns, most crucially – they overlook the interplay of these spreading patterns with those of the pathogens. This results in a discrepancy between supply and demand, that prohibits efficient mitigation even under optimal conditions of superfluous drug/vaccine flow. Therefore, here, we design a dissemination strategy that naturally follows the predicted spreading patterns of the epidemic, optimizing not just for supply volume, but also for its congruency with the anticipated demand. Specifically, we show that epidemics spread relatively uniformly across all destinations, and hence we introduce an equality constraint into our dissemination that prioritizes supply homogeneity. This strategy may, at times, slow down the supply rate in certain locations, however, thanks to its egalitarian nature, which mimics the flow of the viral spread, it provides a dramatic leap in overall mitigation efficiency, saving more lives with orders of magnitude less resources.

---

As international mobility continues to grow, so does the threat of global pandemics^1–6^ as clearly demonstrated in the worldwide inpact of COVID-19^7–12^. Originating in a random outbreak, the pathogens spread internationally though air-travel, then locally, at each destination via contagion dynamics^13,14^. When such scenario transpires, the challenge is to (i) develop *ad hoc* a drug or vaccine; then (ii) design an efficient strategy for its dissemination. The point is that even if a therapeutic is avalibale, eliminating challenge (i), we must still ship it to multiple destinations worldwide – potentially stretching our transportation resources to their maximal capacity. Hence, to outrun the epidemic, we crucially need optimal dissemination schemes, that achieve the most efficient mitigation, given the constraints imposed by our production/shipping limitations^15–19^.

The common approach to this challenge treats the dissemination as a commodity flow problem^20–23^. The therapeutics (commodity) are shipped from one or few sources, propagating along transportation routes – each with a given shipping capacity – aiming to fill the demand of all network nodes. In this framework one seeks the shipping sequence that optimally utilizes the network – namely, allows for the maximal *volume* of commodity to flow through the network per unit time^24,25^ **(Fig. 1a)**. This well-established approach, however, overlooks the interplay between the supply, *i.e*. the therapeutics, and the demand, here generated by the spreading pathogens. *Indeed, our true goal is not just to generate the maximal outflow of commodities, but rather to obtain the optimal mitigation of the disease*. This does not depend only on the *volume* of commodity flow, but also on the manner in which this volume is distributed across all potential destinations, *i.e*. its *spreading pattern*^26,27^. Consequently, the most efficient shipping sequence might be one that compromises maximal flow, but generates the optimal spreading patterns, compatible with those of the pathogens^13,28–32^.

**Figure 1.**
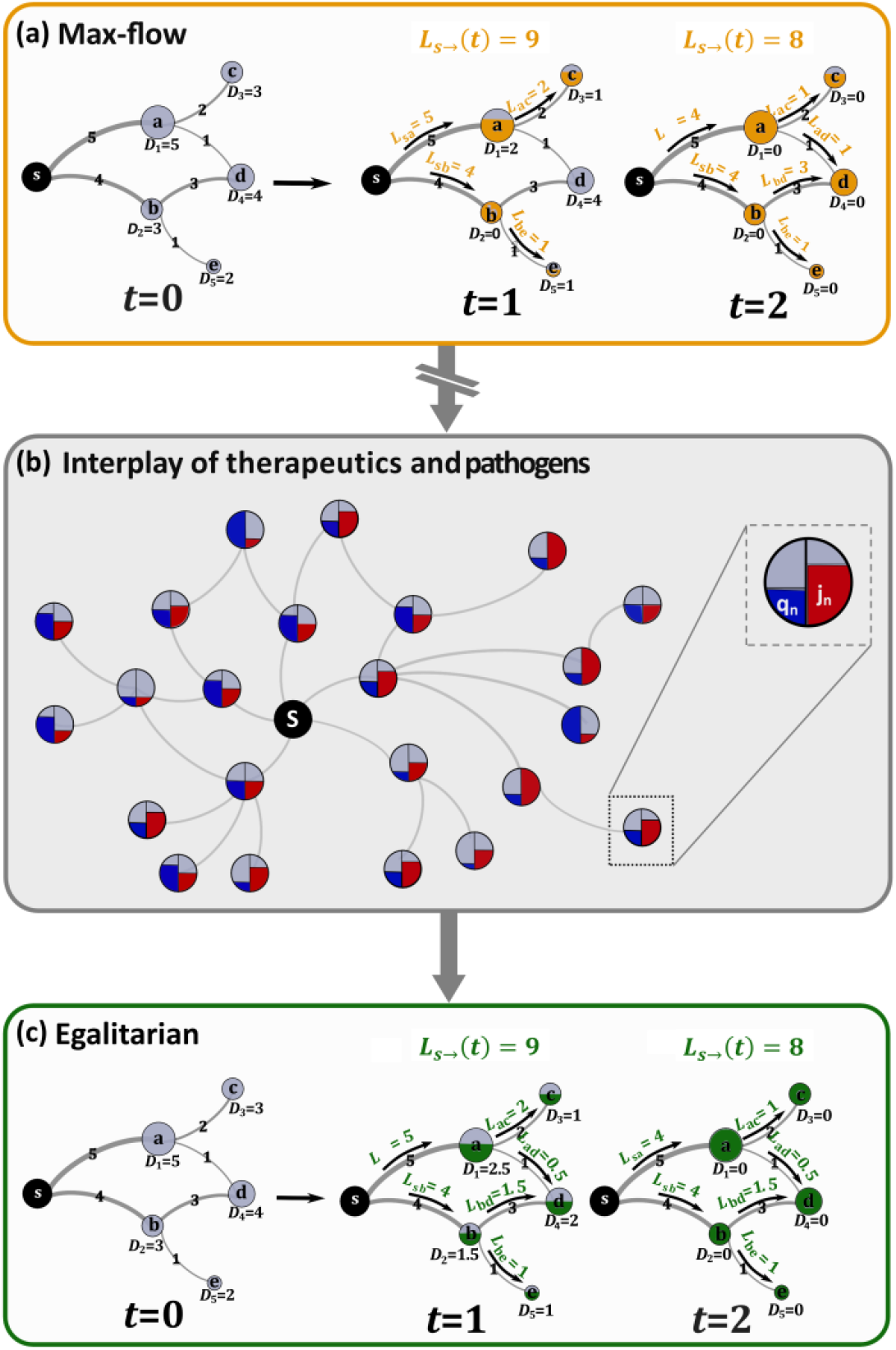
Optmizing therapeutic dissemination for efficient mitigation. We treat the theraputic spread as a commodity flow problem. (a) Commodities flow from the source *s* to fill the demands *D_n_* (*t*) at all destinations *a* through *e*. The transportation network imposes constraints though the restricted shipping capacity of all links. For example, the route *s → a* can transport at most 5 units per day (edge labels). The shipping sequence determines the quotas shipped via each link *L_nm_*, and hence the rates by which all nodes are supplied. Demands are updated as therapeutics are supplied – for instance, at *t* = 1 *b*’s demand *D*_2_(*t*) is updated to zero, as *b*’s demand is filled. Under *max-flow* the goal is to achieve the highest net outflow of commodities from *s, i.e*. maximize *L_s→_*(*t*). Here, nodes were supplies within two days, a net flow L*_s→_*(1) *=* 9 in day 1 and *L_s→_*(*t*) *=* 8, in day 2. (b) During an epidemic, however, the goal is to achieve optimal *mitigation*, not just optimal commodity flow. This requires us to consider the interplay between our dissemination and the spread of the epidemic. Each node’s demand is determined by its infection levels (*j_n_*(*t*), red), leading us to seek the shipping sequence which best adresses the anticiated spread of infections – aiming for supply (*q_n_* (*t*), blue) that is most compatible with demand. (c) We find that the optimal strategy is to design an *egalitarian* flow, in which supply is homogeneously spread across all destinations. Using the same network as in (a), our egalitarian strategy yields an even shipping sequence, in which nodes are supplied concurrently at roughly equal rates. For example, instead of fully supplying node *b* in *t* = 1 and only then shipping to *d* at *t* = 2, as done under max-flow in panel (a), our egalitarian algorithm favors the shipping sequence where *b* and *d* are simulataneasly supplied at equal rates. Under global demand, we show, such egalitarian supply is orders of magnitude more effective in terms of mitigation and resource disposal.

With this in mind, we take here a network dynamics approach, and analyse the spatiotemporal propagation patterns of both the commodities and the pathogens **(Fig. 1b)**. We find that while the disease impacts the majority of nodes uniformly, maximum commodity flow optimization naturally yields a highly uneven spread, conflicting a concurrent and homogeneous global demand with an extremely heterogeneous supply pattern. As a result, not only does maximum flow not guarantee optimal mitigation, it is, in fact, counterproductive, indeed, generating a desirable *volume* of available therapeutics, but at the same time a highly undesirable *spreading pattern*. We, therefore, introduce an *equality* criterion to the optimization, that prioritizes dissemination sequences with homogeneous spreading patterns **(Fig. 1c)**. This ensures that the therapeutic flow is not just rapid, but also properly disseminated, generating supply patterns that are compatible with demand. The resulting dissemination, we find, is orders of magnitude more efficient, achieving higher mitigation levels even under extremely prohibitive shipping constraints.

## MITIGATION VIA MAXIMUM-FLOW

To examine our response to a global epidemic, we consider different epidemiological scenarios, from mildly contagious to extremely virulent, in which a disease spreads globally via air-travel, under the susceptible-infected-recovered (SIR) epidemic model^13,14,33^ **(Box I)**. We used empirical data on human aviation to evaluate the flux of passengers between *N* = 1,292 local populations (nodes), each with *M_n_* individuals (*n* = 1*, …,N*), and quantified the impact of the epidemic through its global *coverage*

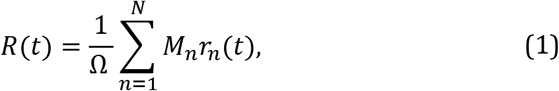

where *r_n_*(*t*) is the fraction of recovered individuals in *n* and 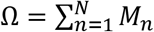 represents the global population. Hence, *R*(*t*) in (1) captures the fraction of impacted individuals worldwide. In **Fig. 2a** we show the unmitigated *R*(*t*) under three different scenarios: extremely contagious, where *R*(*t* → *∞*) → 1 (dark red), medium (*R*(*t* → *∞*) ≈ 0.5, red) and mild (*R*(*t* → *∞*) ≈ 0.3, light red).

**Figure 2.**
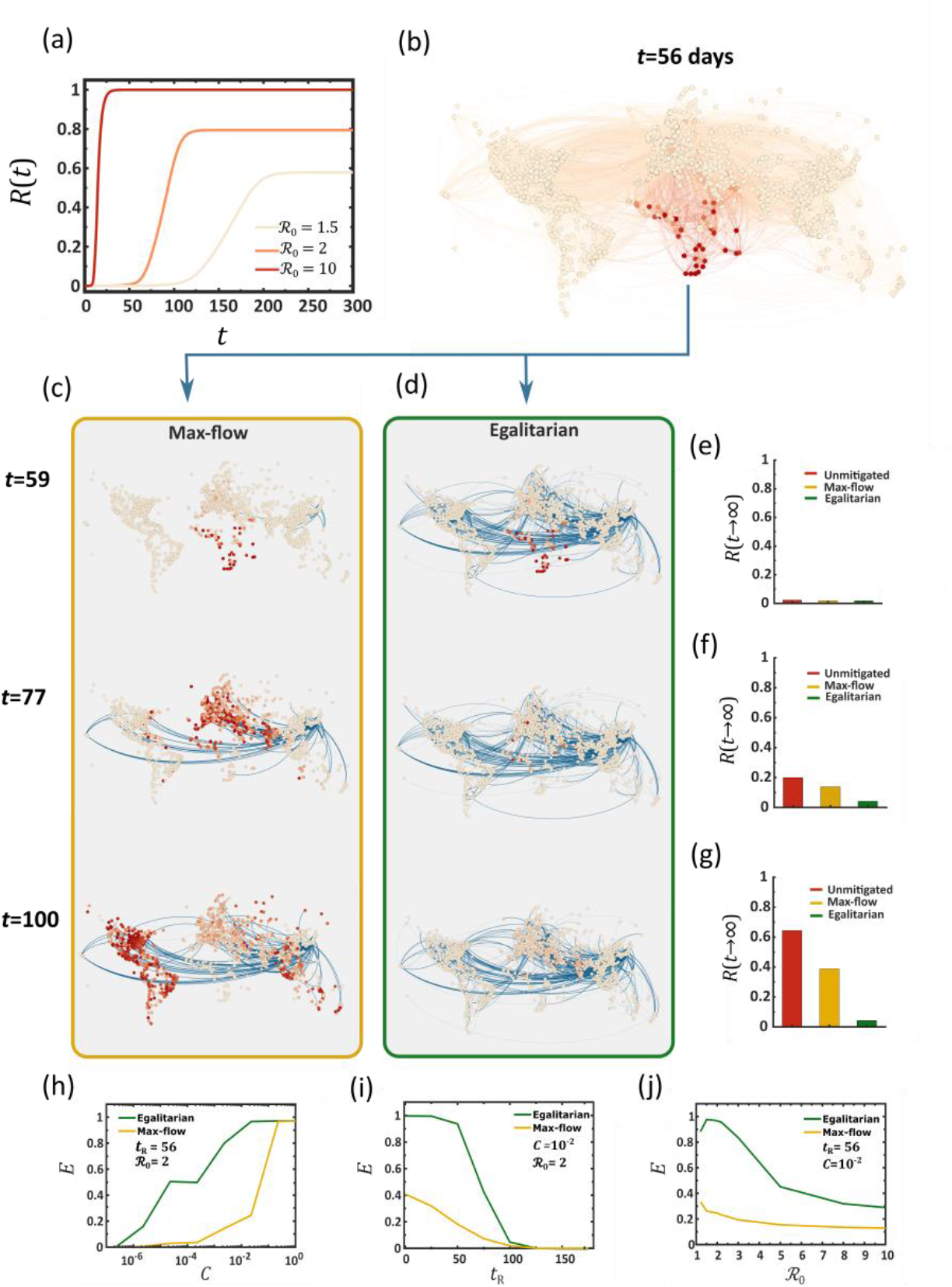
Outrunning a contageuous epidemic using max-flow vs. egalitarian dissemination. (a) The global coverage *R* (*t*) vs. *t* following an outbreak at Burundi (BJM) under three scenarios: severe (dark red ℛ_0_ *=* 10), intermediate (red, ℛ_0_ *=* 2) and mild (light red, ℛ_0_ *=* 1.5). (b) The state of the epidemic at *t* = 56 days, directly before drug dissemination begins. The local coverage *r_n_*(*t*) in each node and the flux of infected individuals along each link (air-route) are represented by their red color depth. (c) *Max-flow dissemination:* starting at *t*_R_ *=* 56 we begin drug dissemination from Osaka (ITM) via max-flow; Eqs. (2)-(5). Drug fluxes *L_nm_*(*t*) (links) and drug availability *q_n_*(*t*) (nodes circumference) are represented by blue color depth. We observe a *race* between the therapeutic and the disease, both spreading along similar routes, ending in a significant fraction of infected individuals, as indicated by the prevalence of red nodes at *t* = 100. (d) *Egalitarian dissemination:* under the same scenario, the egalitarian strategy of Eqs. (11)-(13) achieves a much more efficient mitigation, where by *t* = 100, the epidemic is almost eliminated. (e)-(g) *R*(*t*) at the three timepoints: unmitigated (red), under max-flow (yellow) and under egalitarian (green). By *t* = 100 egalitarian has reduced the global coverage by a factor of ~ 20, from *R*(*t*) *=* 0.65 to *R*(*t*) *=* 0.04, while max-flow has achieved a mere ~ 1.5 factor reduction. Three crucial parameters may impact our mitigation: the global transportation capacity *C*, our response time *t*_R_ and the severity of the contagion ℛ_0_. (h) Mitigation efficiency *E* vs. *C* under egalitarian (green) and max-flow (yellow). The former achieves higher efficiency with orders of magnitude less resources. For example, egalitarian provides *E* = 0.8 under *C ~* 10^−3^, as compared to *C ~* 10^−1^, a two order of magnitude gap, required under max-flow for similar efficiency. (i) *E* vs. *t_R_*. Again, we find that egalitarian shows a significantly higher performance in the face of a late response. (j) *E* vs. ℛ_0_, confirming, again, the consistent advantage of egalitarian mitigation. In each panel we vary one parameter and list the set values of the other two.

Following the initial outbreak at *t* = 0, we define the response time *t*_R_ as the time required to begin the distribution of a therapy. This therapy can be in the form of a vaccine, designed to prevent the infection of *susceptible* individuals, or a drug, facilitating the recovery of *infected* individuals. As we consider a scenario in which the disease is rapidly spreading, and hence many individuals may already be infected, we focus below on the dissemination of drugs, which remain relevant at all stages of the spread^34,35^.

Examining the efficiency of *dissemination*, rather than production, we assume that the therapeutic is already stockpiled in sufficient quantities at a specific source node *s*. Therefore, the main challenge is to optimally distribute it via the air-transportation network to all destinations. The network is characterized by the *carrying capacities B_nm_* that quantify the daily volume of drugs that can be shipped through each air-route *m → n, i.e. B_nm_ = B_n←m_*. The challenge is, therefore, to obtain the optimal shipping sequence from s to all other nodes, under the constraints imposed by *B_nm_*.

Max-flow mitigation (Fig. 2a-c).

In the maximum flow dissemination strategy we seek to maximize the daily volume of drug doses spreading from s to the rest of the network. Denoting the number of doses shipped from *m* to *n* on day *t* by *L_nm_*(*t*), our optimization translates to

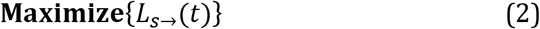

where

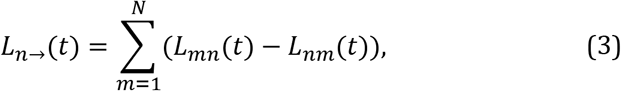

captures the net daily outflow of drugs from *n*. Hence, in Eq. (2), by maximizing *L_s→_*(*t*)*, i.e*. the outflow from the source, we seek to enhance the total volume of drugs introduced into the network in each day. This maximization is subject to two constraints:

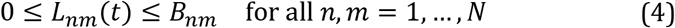

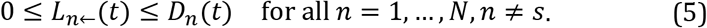

The first constraint (4) ensures that the daily flux along each route *m → n* is within the bounds of the route carrying capacity *B_nm_*. The second constraint (5) restricts nodes from accumulating doses in excess of their remaining demand at time *t*. Hence *n’s* net *incoming* flux *L_n←_*(*t*) is bounded by *n’s* current demand, *D_n_*(*t*); note that *L_n←_*(*t*) *= −L_n→_*(*t*).

We consider three strategies for setting the demands: (i) *Population based*. At *t* = 0 we set *D_n_*(0) to be proportional to each node’s population *M_n_*, then update these demands daily according to *n’s* supply, as *D_n_*(*t* + l) = *D_n_*(*t*) *− L_n←_*(*t*) • (ii) *Impact based*. Setting *D_n_*(0) according to the projected number of infected individuals at *n* • (iii) *Urgency based*. Updating *D_n_*(*t +* 1) dynamically based on *n’s* supply gap at *t*, such that highly impacted (or undersupplied) nodes are prioritized. While (i) is simplest, it is also inefficient, tending to over-estimate the actual demand at *n;* (iii), on the other hand, is highly complex, relying on real-time tracking and updating of the demands, but, in some cases, benefits, by design, from a high level of congruency with the spread of the pathogens. An expanded discussion of these three strategies appears in **Supplementary Section 2.4**.

**Network capacity**. Applying (2) – (5), the maximal dissemination rate one can achieve is restricted by the out-degree of the source node, 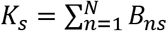, representing an upper-bound on the volume of drugs that can be introduced into the network per day. We, therefore, define the network’s normalized *capacity* with respect to each source *s* as

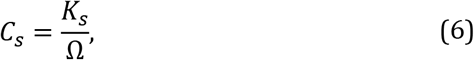

normalized to *C_s_* = 1 in case *s* can disseminate sufficient doses to meet the entire global demand (Ω) in one day. The network capacity can be controlled by rescaling all individual carrying capacities *B_nm_*, to describe affluent vs. restrictive dissemination scenarios **(Supplementary Section 2.3)**.

In **Fig. 2b,c** we present the evolution of the epidemic at four selected time-points. At *t* = 0 we simulate an outbreak (red) at Burundi (BJM), emulating the 2013 Ebola, which originated in Africa^36,37^, then track its spread through air-travel, setting ℛ_0_ *=* 2. The node infection levels and the epidemic fluxes, *i.e*. the daily volume of infected passengers on each route, are represented by red color depth. Drug dissemination via max-flow optimization (blue) begins at *t*_R_ *=* 57 days in Osaka (ITM), using blue color depth to signify the availability/flux of drugs in each node/route. We set the network capacity in Eq. (6) to *C* = 10*^−^*^2^, a dissemination capability of 1% of the global demand per day, and assign demands *D_n_*(*t*) using the impact based strategy. We find, through the long-term prevalence of infections (**Fig. 2c**, red), that under these conditions, mitigation falls short. Indeed, in **Fig. 2e-g** we observe that the coverage *R*(*t*) in (1) is only slightly affected by our max-flow mitigation (red vs. yellow), illustrating the failure to effectively suppress the epidemic.

For a more systematic assessment of our dissemination strategy, we track the mitigation efficiency via^15^

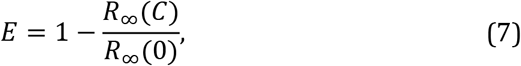

where *R_∞_*(*C*) is the long term coverage, *i.e. R*(*t* → *∞*), of the epidemic under mitigation with network capacity *C_s_ = C*. In case mitigation is effective we expect *R_∞_*(*C*) ≪ *R_∞_*(0), a significant reduction in the disease coverage, which translates to *E →* 1. Conversely, a failed mitigation leaves infection levels almost unchanged, leading in (7) to *E →* 0. Testing *E* vs. *C* under maximum flow dissemination, we find that for a broad range of *C* levels the epidemic is almost unaffected, a consistently inefficient mitigation in which *E* ≪ 1 (**Fig. 1h**, yellow). Effective mitigation is only achieved around *C* ≳ 0.1, a limit in which s is capable of shipping doses in the order of the entire global demand in few days. Such optimal conditions are not only unlikely, but mainly, they indicate the inefficiency of this dissemination strategy, requiring an extreme volume of therapeutics shipped in a highly constrained timeframe in order to achieve a measurable impact on the epidemic.

It seems, therefore, that the max-flow optimization strategy is inadequate for the containment of a globally spreading epidemic. In **Supplementary Section 2** we analyze the population and urgency based strategies, observing similar challenges.

### The roots of the max-flow inefficiency

Our mitigation is, in its essence, a spreading competition between the therapeutics and the pathogens, both progressing along the same underlying network, *i.e*. air-transportation^19^. It seems, therefore, that winning this competition is a matter of shipping capacity: we must generate sufficient therapeutic fluxes to outrun the spread of the disease, namely we must increase *C_s_*. However, the analysis above indicates, that there is an intrinsic deficiency in the spread of therapeutics, that cannot be easily compensated by simply increasing shipping rates. Next, we show that the two competing processes – viruses vs. therapeutics – indeed, lead to fundamentally different spreading patterns, in which the viruses benefit from an intrinsic advantage.

**Propagation of pathogens (Fig. 3a-c)**. Viruses spread via diffusion coupled with local SIR dynamics, as captured by Eqs. (15) in **Box I**. In this process, upon penetration of node *n*, the viruses reproduce locally through SIR, until reaching peak infection at 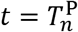. In a network environment, since the majority of nodes are at the mean distance from the initial outbreak, we find that after a limited propagation time, most nodes reach peak infection approximately simultaneously. We track this via the infection rate

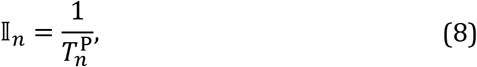

capturing the average *speed* by which the pathogens propagate to *n*. In **Fig. 3c** we show 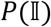, the probability density for a randomly selected node *n* to have 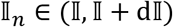. Indeed, we find that 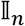 follows a bounded distribution, where the majority of nodes are infected at a similar rate, and hence their peak infection occurs at roughly at the same time. Such uniform propagation patterns lead to a simultaneous global peak infection, and, as a consequence, to a concurrent global demand for therapeutics.

**Figure 3.**
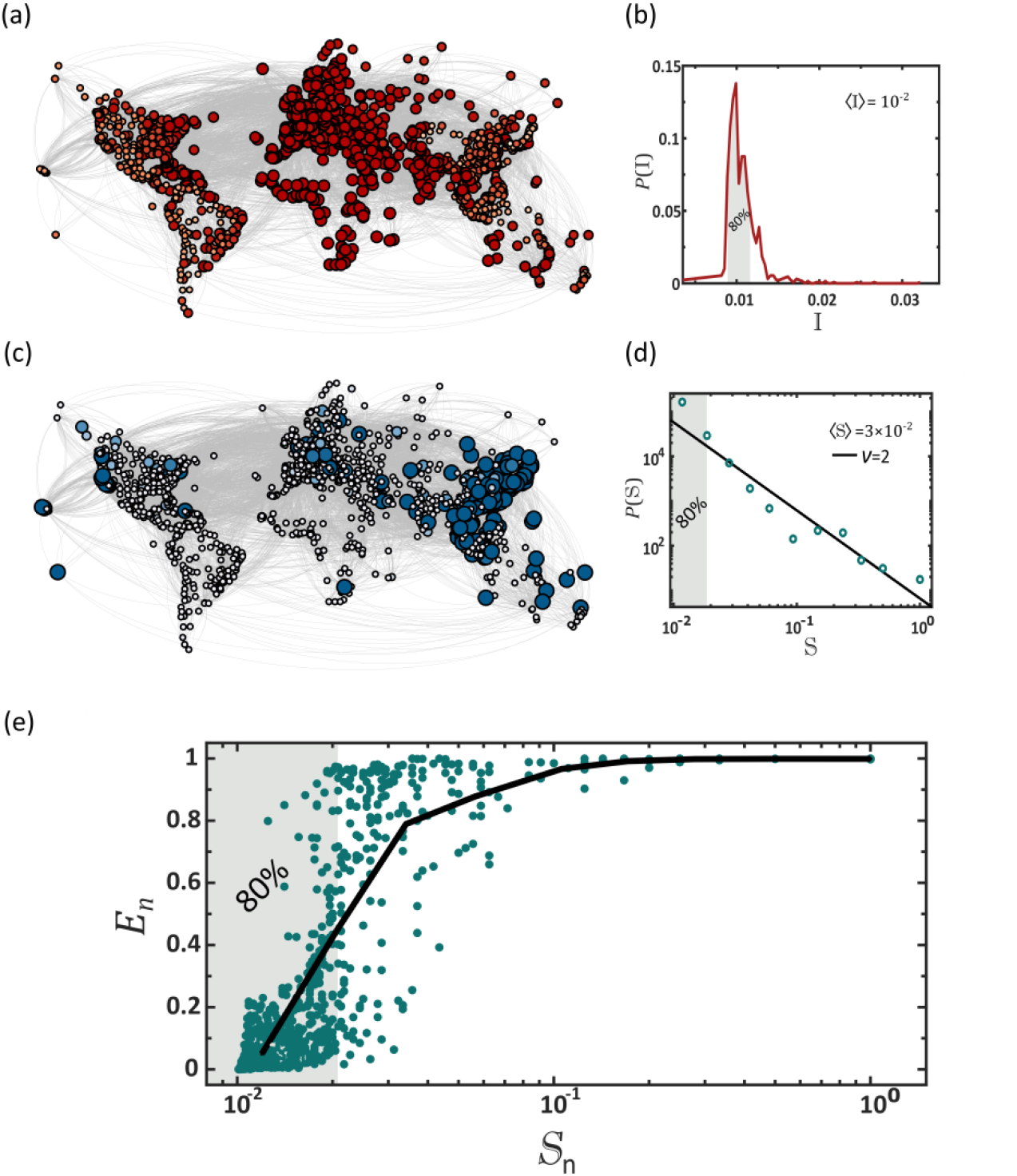
Why max-flow provides sub-optimal mitigation. (a) Pathogens spread via contagion dynamics, reproducing in each desination independently via the SIR process. The result is a relatively homogeneous spread, in which the majority of destinations reach peak infection (red) at approximately the same time. Here this is observed by the almost uniform red shade and size of all nodes, capturing the instantaneuous infection level *j_n_*(*t*) during the time of global peak infection. (b) To quantify this we measure the infection rate 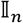 of all nodes as obtained from Eq. (8), and plot its distribution 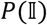. The bounded form of P(I) indicates the homogeneous nature of the pathogen spread. Indeed, we find that 80% of the nodes are impacted wthin a small margin around the average (shaded area). (c) Using max-flow we track the availability of the therapeutics *q_n_*(*t*) at all destinations (node size/blue color depth). In contrast to the pathogens, the therapeutics naturally spread extremely unevenly, with a small minority of early supplied nodes (large, dark blue), coexisting alogside a majority of delayed destinations (small, light blue). (d) The supply rate distribution 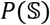 as obtained via max-flow (circles). As predicted 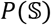 can be approximated by a power-law 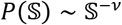, with *ν* = 2 (solid line). This captures an extremely heterogeneous dissemination pattern, in which 80% of the nodes are supplied at a below average rate (shaded area). Hence, max-flow confronts a homogeneous demand (*j_n_*(*t*), red) with an extremely uneven supply (*q_n_*(*t*), blue). (e) The local efficiency *E_n_* vs. supply rate 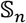 as obtained via max-flow dissemination. As expected we find that 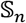 crucially impacts the mitgation effectiveness at *n*. Therefore, the fat-tailed nature of 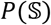, in which the majority of nodes have a small 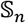 (80%, shaded) creates an intrinsic mitigation defficiency, in which most destinations experience insufficient supply. The challenge, we emphasize, is not in the production/shipping capacities, as indeed we have 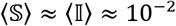, namely, that on average, supply rate can meet the evolving demand. Rather, is is rooted in the patterns of spread of the existing supply, which are fundamentally incongruent with those of the pathogens.

**Dissemination of therapeutics (Fig. 3d-f, Supplementary Section 4)**. The dynamics of commodity flow, as provided by Eqs. (2) – (5) are fundamentally different. The therapeutics flow from a single source node *s*, and undergo dilution as they disperse across the exponentially growing number of pathways^38,39^. Such flow patterns naturally lead to a highly heterogeneous supply pattern across all nodes. To observe this we consider the therapeutic supply time 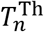 for node *n* to fill its therapeutic demand, and its consequent supply *rate*

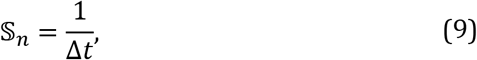

where 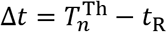, is the elapsed time from initial dissemination (*t*_R_) to *n*’s final supply 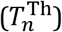. This rate 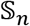 quantifies how rapidly *n* receives treatment. In **Supplementary Section 4** we show that under max-flow mitigation the probability density 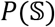 follows a power law of the form 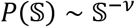, with *ν* = 2 **(Fig. 3f)**. Hence, in contrast to the homogeneity of the epidemic spread, *supply* is extremely heterogeneous, with a vast majority of undersupplied nodes (small 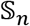, shaded 80%), and a selected privileged minority of well-treated destinations (large 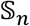).

Together, this combination of *homogeneous* demand and *heterogeneous* supply, a consequence of the naturally occurring spreading patterns of pathogens versus commodities, creates a crucial gap in our ability to effectively mitigate a global pandemic. To understand this, consider ideal flow conditions, where 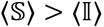, namely that, on average, dissemination is more rapid than the viral propagation. Still, the fat-tailed nature of 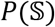 vs. the bounded form of 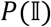 indicates that while most nodes have 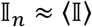, when it comes to supply, the typical node has 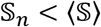. Indeed, in a scale-free distribution the majority of entries are below the mean. This is illustrated in **Fig. 3**, where we show that 80% of nodes are within a 20*%* margin of 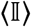, *i.e*. they are all impacted within a narrow time window around the mean (**Fig. 3b**, shaded). Yet, in contrast, a similar 80% have a therapeutic rate below 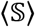, *i.e*. they witness supply deficiency (**Fig. 3d**, shaded). Hence we are confronted with a reality in which almost all nodes require treatment, and yet only a small minority receives sufficient, and at times, even superfluous, supply.

This uneven distribution 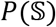 directly impacts our mitigation efficiency. To observe this, consider the local efficiency

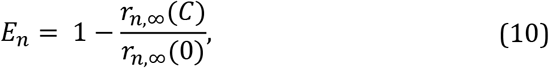

where *r_n,∞_*(*C*) *= r_n_*(*t* → ∞) under mitigation with capacity *C_s_ = C*. Hence, while *E* in (7) captures the global mitigation efficiency, *E_n_* focuses specifically on the effect observed at *n*. In **Fig. 3e** we show *E_n_* vs. 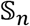. As predicted, we observe that 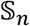 is a crucial determinant of mitigation efficiency. Indeed, nodes with low 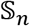 (shaded), by far the majority of nodes according to the power-law structure of 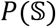, exhibit an almost vanishing *E_n_*. This illustrates, once again, the flaw of max-flow: most nodes, the deprived 80%, have *E_n_ →* 0, while a small minority benefit from *E_n_ →* 1.

Taken together, our analysis shows that the common approach of maximizing flow is insufficient, as it treats the *mean* flow, but not its *distribution*. Most essentially, it disregards the interplay with the competing flow of the pathogens. When confronting these two spreading processes, we observe that the main challenge is rooted in equality 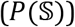 rather than in quantity 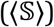, namely not the volume of the outflux, *L_s→_*(*t*), but rather the way this volume distributes across the network. Hence, below we complement our optimization for max-flow with an additional requirement, seeking supply homogeneity.

## OPTIMIZING FOR HOMOGENEITY

**Egalitarian mitigation (Fig. 1c)**. The optimization of Eqs. (2) – (5) is designed to increase the volume of drugs introduced into the network, but as we have shown, it leads to an extremely sub-optimal dispersion pattern. To remedy this, we introduce a homogeneity criterion, replacing the maximization in (2) by (**Supplementary Section 2**)

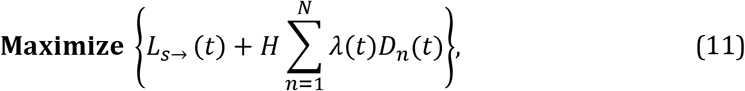

where the homogeneity coefficient *H* determines the relative balance between our optimization for max-flow (*L_s→_*(*t*)) vs. our demand for egalitarian spread (*λ*(*t*)). In addition to constrains (4) and (5) above, we now also require

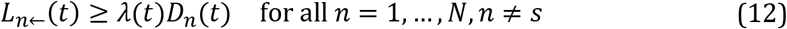

and

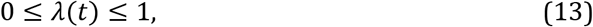

forcing, at each step, to supply, at the least, a fraction *λ*(*t*) of every node’s current demand. This strategy seeks to maximize flow, but at the same time favors solutions that supply all nodes simultaneously. Indeed, constraint (12) prohibits shipping sequences that at any step supply exclusively a subset of the nodes. This additional constraint may, in general, lead to a diminished net flow, however, thanks to its egalitarian nature, we find that it dramatically enhances mitigation efficiency.

To observe this, we revisit the epidemic spread of **Fig. 2**, leaving all conditions unchanged, except that now, instead of the max-flow optimization of (2) – (5) we mitigate the disease via the egalitarian strategy of (11) – (13). The results are striking: egalitarian drug dissemination practically eliminates the epidemic, as observed by the dominance of blue nodes/links at *t > t*_R_ (**Fig. 2d**). Indeed, compared to the ~ 30*%* reduction in *R*(*t* → ∞) afforded by max-flow, egalitarian achieves, under the same capacity *C_s_*, a ~ 95*°%* reduction, representing a practically perfect mitigation **(Fig. 2g)**.

A systematic comparison of the two mitigation strategies consistently supports the crucial role of dissemination homogeneity. In **Fig. 2h** we show the efficiency *E* in (7) vs. *C* under egalitarian dissemination (green). We find that *E* approaches 80% already at *C ~* 10^−3^, representing an extreme case of supply deficiency. A similar *E* under max-flow was only observed for *C_s_ ~* 10^−1^ (yellow), a two order of magnitude advantage for egalitarian. A crucial factor impacting our mitigation efficiency, is the response time *t*_R_, required to identify the threat and initiate a response. To observe this, in **Fig. 2i** we present *E* vs. *t*_R_ for both max-flow (yellow) and egalitarian (green) mitigation. As expected, we find that *E* declines with *t*_R_, however, for the entire range of response times egalitarian consistently outperforms max-flow. In **Fig. 2j** we further show that the egalitarian advantage is consistently maintained for a range of contagion levels ℛ_0_.

Hence, examined against a series of potential challenges, from the disease parameters (ℛ_0_) to the performance level of our response (t_R_, *C_s_*), we find that egalitarian allows enhanced mitigation, in some cases, by a dramatic margin. This, again, is thanks to egalitarian’s homogeneous dissemination patterns, which, under the price of a potentially diminished supply *rate*, afford us a more desirable supply *distribution*, and consequently, an optimized mitigation.

### The roots of the egalitarian advantage

The efficiency of egalitarian mitigation may seem, at first glance, implausibly high. For example, **Fig. 2h** indicates that effective mitigation is already achieved with capacity as low as *C* ~ 10^−3^ day^-1^. This represents an extreme scenario, in which we require an order of 10^3^ days to supply the global demand, a time scale that by far exceeds the final spread of the epidemic. Such extraordinary efficiency is rooted in the fact that even a tiny influx of drugs introduced at an early enough stage of the epidemic can dramatically impact its future development.

To understand this, consider a node *n* at the early stage of the spread, when *j_n_*(*t*) ≪ 1. As time advances, this small seed of infections reproduces until *n* reaches its final infection levels *r_n_*(*t → ∞*). Suppressing this seed when it is small is, therefore, the optimal strategy, allowing us, with a tiny supply of therapeutics, to eliminate not just the current *j_n_*(*t*), but also terminate its potential future reproduction. Hence, maintaining a continuous influx of therapeutics, even at a very low rate, keeps *j_n_*(*t*) subdued, never allowing the local infections to reach their potential coverage. Egalitarian mitigation is precisely designed to provide such supply patterns: instead of rapidly supplying a select group of nodes at a time, it favors a slow influx that is spread evenly across all nodes, starting from the *get-go* at *t = t*_R_.

In contrast, max-flow generates a similar (or even higher) total flux, but spreads it out sequentially – few nodes receive all their supply early on, and are hence successfully treated, while the majority fill their demand at a much later time, when *j_n_*(*t*) is already harder to contain. To observe this, we revisit the simulated mitigation of **Fig. 2**, this time focusing on a pair of specific nodes **(Fig. 4a)**: SDJ which is among the few early supplied nodes (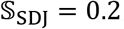, red), and CGQ, a typical node, which represents the majority destinations, which receive their supply later on (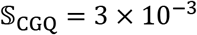, Color). In **Fig. 4b,c** we show the drug availability *q_n_*(*t*) in both nodes (blue), together with their unmitigated (red) vs. mitigated (yellow) disease coverage. Indeed, we find that early supply is crucial, with SDJ benefiting from an almost perfect mitigation, and CGQ being almost unaffected by the late arriving drugs.

**Figure 4.**
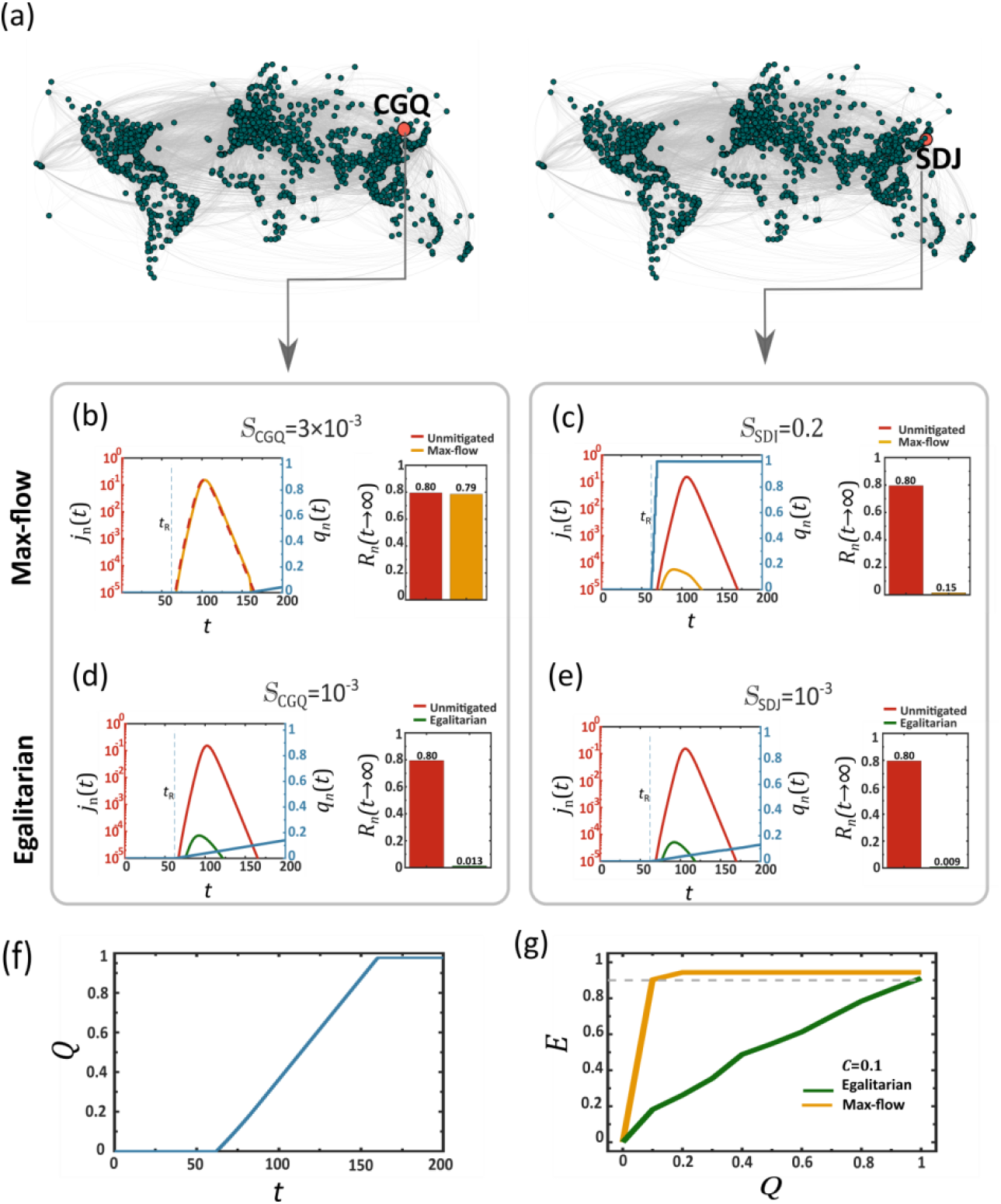
The roots of the egalitarian advantage. (a) We focus on two specific nodes, SDJ (right), who, under max-flow, is among the early supplied nodes 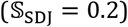, and on CGQ (left), representing the majority of undersupplied destinations 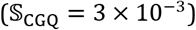. (b) Therapeutic availability *q_n_*(*t*) vs. *t* at CGQ (blue) as obtained under max-flow dissemination. Due to the sequential nature of this dissemination scheme, the therapeutic reaches CGQ too late, and consequently its infection level *j_n_*(*t*) remains high, both without (red) and under the effect of max-flow mitigation (yellow). (c) In contrast, SDJ is among the first to fill its demand, hence the sharp rise in *q_n_*(*t*) at an early stage (blue). As a result this node benefits from highly efficienct mitigation, with *r_n_*(*t → ∞*) reduced dramatically from 80% (unmitigated, red) to a mere 15% (mitigated, yellow). The challenge is that while CGQ is representative of the majority of destinations, the highly benefited SDJ belongs to a select minoroty under the max-flow framework. (d)-(e) Egalitarian generates a homogeneous supply pattern, hence now both CGQ and SDJ share a similar (low) supply rate of ~ 10^−3^. Despite that they both benefit from a near perfect mitigation (unmitigated – red; mitigated – green). This is thanks to the fact that despite their *equal but slow* supply, they now both begin to receive the therapeutic at an early stage (*q_n_*(*t*), blue). Indeed, even SDJ, who is now, under egalitarian, experiencing a dramtic drop in its supply rate, from 0.2 to 10^−3^, continues to benefit from highly efficient mitigation. Hence, the simulataneous, albeit uniformly slow, treatment afforded via egalitarian, not only benefits the max-flow deprived majority, but also continues to effectively treat its superflous minority. (f) The global consumption *Q*(*t*) vs. *t*, approaching unity, *i.e*. all demands are filled, when *t → ∞*. (g) Global mitigation efficiency *E* vs. the volume of disseminated therapeutics *Q*. Under max-flow (yellow) efficient mitigation *E →* 1 requires *Q →* 1 (yellow). In contrast, egalitarian has *E ~* 1 already at *Q ~* 0.1, achieving, thanks to its concurrent supply patterns, an almost perfect mitigation with only 10% of the resources.

Under egalitarian mitigation the supply pattern is fundamentally different: indeed, as per the equality constraint, both nodes settle for the slower rate 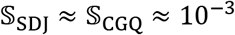. Hence, egalitarian provides no overall rate advantage, and, in fact, disadvantages the high rate SDJ. However, now, instead of *q_n_*(*t*) remaining idle and then, in due time, receiving a sharp supply boost, it is continuously supplied from the outset at *t_R_* **(Fig. 4 d,e)**, a direct consequence of the homogeneity constraint of (11) – (13). The advantage is that now both SDJ and CGQ receive treatment early on, mitigating the disease while it is still small, and thus suppressing its potential to grow. Therefore, while egalitarian impedes the supply of the high rate minority, it still allows us to successfully treat both that minority and the low rate majority. This is because its simultaneous, even if slow, supply, affords early treatment for *all* nodes, overcoming *j_n_*(*t*) while it is still at its embryonic stage.

### Mitigation under restricted supply

A crucial implication of the above analysis touches on the global resources required for mitigation^40,41^. Clearly, running the max-flow/egalitarian algorithm until all demands are filled will inevitably cost a total of ~ Ω drug units, supplied at a time-scale of ~ Ω*/C_s_* days. However, since egalitarian begins supplying all nodes while *j_n_*(*t*) is still small, it has the potential to overcome the pandemic even before all initial demands *D_n_*(0) are completely satiated. To observe this, we run both strategies – max-flow and egalitarian – for a limited time, from *t* = 0 to *T*. We then measured the efficiency *E* (7), in function of the total resource consumption

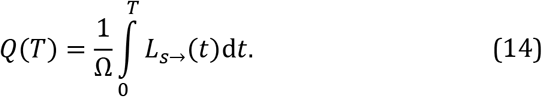

Equation (14) quantifies the total outflux of therapeutics from *s* as a fraction of the global demand Ω, throughout the mitigation period, *t* = 0 to *T*. The greater is *Q*(*T*), the more therapeutic resources that are consumed. As predicted we find in **Fig. 4g** that egalitarian, not only provides a higher mitigation efficiency, but achieves this in a fraction of the resources. For max-flow (green) we observe a linear gain with *Q*, reaching peak efficiency only around *Q* → 1*, i.e*. when supply fills *all* demands. Egalitarian, in contrast, thanks to its simultaneous treatment of all nodes, suppresses the epidemic at an early stage, when it is still manageable, with limited supply, therefore enabling mitigation of above 90*%* (grey dashed line) with total consumption as low as *Q ~* 0.1, *i.e*. a mere 10*%* of the global demand.

## DISCUSSION

Commodity flow problems are at the heart of many crucial applications, from communications to supply chains, seeking to optimize a network’s capacity to distribute information or goods^20-25,42-44^. Most often the target optimization function, *e.g*., max-flow, arises naturally from the purpose of the distribution. However, in the context of disease mitigation, the optimization is a consequence not just of the dissemination scheme, but also of its interplay with the viral spread. Indeed, our true goal is not just distribution efficiency in and of itself, but rather *mitigation* efficiency. Hence, to assess a dissemination protocol we must couple it with the spread of the epidemic, and observe its effectiveness in terms of the actual observed reduction in infection levels. Here we have shown that the naïve approach of maximizing the commodity flow fails this test, indeed, providing rapid supply, but at the same time a highly inefficient mitigation. On the other hand – optimizing for homogeneity dramatically improves mitigation efficiency.

Our findings are relevant in case the epidemic spreads globally – a scenario averted in many recent pandemics, but experienced quite severely during COVID-19. In such scenario, the peak infection occurs approximately simultaneously at logarithmic time-scales, due to the short network paths^27,45^. Our egalitarian protocol, by spreading simultaneously, albeit slowly, to all nodes outlines the most effective strategy, catching the disease at its early stage in all locations. To fully benefit from this advantage, it is crucial to activate the egalitarian dissemination as early as possible, to truly attack the epidemic at its seed. Otherwise, social distancing measures must be employed to delay the viral spread and keep it at manageable levels until egalitarian dissemination can be instigated^46–52^.

In this regard, the recent successes in epidemic forecasting^53^ are key: first, predict whether the pathogens *will* spread globally, then instigate egalitarian dissemination. Of course, a false alarm might result in a waste of resources, however our analysis indicates that early intervention allows mitigation with relatively small amount of doses, hence we believe that the potential gain by far outweighs the risk.

Battling a global spread, it is crucial to appropriately set the demands *D_n_*(*t*), which determine how much supply will be shipped to each node. On the side of caution, the natural tendency is to overestimate these demands, *i.e*. generate redundancy. However, in the face of a virulent epidemic, where the distribution network is stretched to its maximal capacity, such strategy may over-allocate resources to some destinations, while depriving others. On the other hand, fine-tuning the demands to meet the exact supply gap of each node, as we do, for example in our *urgency-based* strategy, is highly complex, not always practical, and, if inaccurate, may lead to undersupply in some destinations. Our egalitarian optimization addresses this trade-off quite naturally, by simultaneously supplying all nodes. To understand this consider two nodes *n* and *m*. Under max-flow, supply will be typically sequential, *e.g., n* and then *m*. Under these conditions, overestimating *D_n_*(*t*) will lead to the shipment of more drugs to *n*, thus wrongly delaying the treatment of *m*. In contrast, such discrepancy under egalitarian, will have no bearing on *m*, as both nodes are simultaneously supplied from the start. This makes egalitarian highly robust against the specific assignment of *D_n_*(*t*), allowing us to design a simple dissemination protocol, free of the need to accurately tailor and fine-tune all demands.

Our analysis focuses solely on the mitigation efficiency, yet its conclusions touch upon the ethics of resource allocation – here, prioritizing the even distribution of drugs in a global emergency. While often one is confronted with a clash of values in such cases, here we find it encouraging that efficiency and equality are, in fact, compatible – representing a case in which scientific results go hand in hand with moral directives.

## Data Availability

Upon publication, the authors plan to make all datasets and numerical codes publicly available.

## Author contribution

All authors designed and conducted the research. AH analyzed the data and conducted the numerical simulations, AH, BB and RC lead the analytical derivations, and IB and SE advised on the biological applicability. BB was the lead writer.

## ADDITIONAL INFORMATION

**Competing interests**. Authors declare no competing interests.

**Data availability**. Upon publication, the authors plan to make all datasets and numerical codes publicly available.

### Box I. Modeling the mitigation of a global epidemic

In a network of *N* coupled nodes *n* = 1*, …,N*, each with a population of *M_n_* individuals, we use the SIR model to track the fraction of *M_n_* who are susceptible (*s_n_*), infected (*j_n_*) or removed (*r_n_*), where the infected are divided as 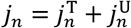, among the treated (T) individuals, who have been provided a therapeutic and the untreated (U) individuals, who have not yet gained access to it. The epidemic dynamics is driven by (**Supp. Sec. 1**)

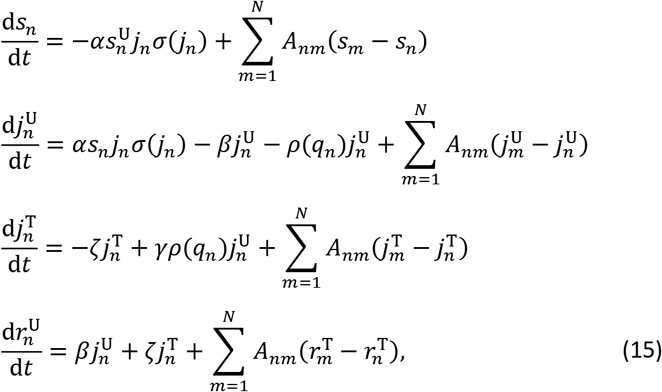

where *α* is the infection rate, *β* is the mortality/recovery rate of the untreated individuals and ζ > *β* is the recovery rate under treatment. The epidemic reproduction number ℛ_0_ = *α/β* captures the level of contagion of the (untreated) disease. The therapeutic consumption rate *ρ*(*q_n_*) depends on the availability of the therapeutic *q_n_*(*t*) in *n*, as

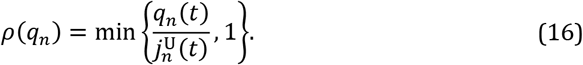

Hence *ρ*(*q_n_*) increases linearly with *q_n_*(*t*) as long as the demand (denominator) exceeds the supply, and saturates to unity when *n* has excess quantities of the therapeutic, avoiding over consumption (**Supp. Sec. 1.2**). The availability *q_n_*(*t*) is determined by our dissemination scheme, following either max-flow, as in Eq. (3), or egalitarian flow, as in Eq. (11).

In (15) we introduce an invasion threshold *ε* through the sigmoidal function

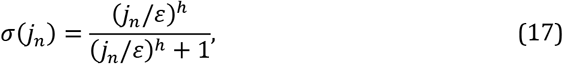

which activates the local SIR dynamics only when the local infection levels 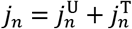 exceed *ε*. The diffusion of individuals between nodes is mediated by *A_nm_*, derived from the empirical international air-travel network (**Supp. Sec. 2.1**).

